# The influence of pH on SARS-CoV-2 infection and COVID-19 severity

**DOI:** 10.1101/2020.09.10.20179135

**Authors:** Leandro Jimenez, Ana Campos Codo, Vanderson de Souza Sampaio, Antonio E.R. Oliveira, Lucas Kaoru Kobo Ferreira, Gustavo Gastão Davanzo, Lauar de Brito Monteiro, João Victor Virgilio-da-Silva, Mayla Gabriela Silva Borba, Gabriela Fabiano de Souza, Nathalia Zini, Flora de Andrade Gandolfi, Stéfanie Primon Murano, José Luiz Proença-Modena, Fernando Almeida Val, Gisely Cardoso Melo, Wuelton Marcelo Monteiro, Maurício Lacerda Nogueira, Marcus Vinícius Guimarães Lacerda, Pedro M. Moraes-Vieira, Helder I Nakaya

## Abstract

The severe acute respiratory syndrome coronavirus 2 (SARS-CoV-2) can infect a broad range of human tissues by using the host receptor angiotensin-converting enzyme 2 (ACE2). Individuals with comorbidities associated with severe COVID-19 display higher levels of *ACE2* in the lungs compared to those without comorbidities, and conditions such as cell stress, elevated glucose levels and hypoxia may also increase the expression of *ACE2*. Here we showed that patients with Barrett’s esophagus (BE) have a higher expression of *ACE2* in BE tissues compared to normal squamous esophagus, and that the lower pH associated with BE may drive this increase in expression. Human primary monocytes cultured in reduced pH displayed increased *ACE2* expression and viral load upon SARS-CoV-2 infection. We also showed in two independent cohorts of COVID-19 patients that previous use of proton pump inhibitors is associated with 2- to 3-fold higher risk of death compared to those not using the drugs. Our work suggests that pH has a great influence on SARS-CoV-2 Infection and COVID-19 severity.

## Introduction

As of August 2020, the severe acute respiratory syndrome coronavirus 2 (SARS-CoV-2) infected over 20 million people worldwide (World Health Organization). The new coronavirus disease 2019 (COVID-19) caused by SARS-CoV-2 is characterized by a broad range of symptoms, from respiratory to neurological and digestive problems (*1, 2*). Although a small fraction of patients develop highly lethal pneumonia, at least 20% of COVID-19 patients may display one or more gastrointestinal (GI) symptoms (*1*), such as diarrhea, vomiting, and abdominal pain (*2, 3*).

SARS-CoV-2 tissue tropism can be directly linked to the diverse clinical manifestations of COVID-19. The receptor used by the virus to enter the cells is the angiotensin-converting enzyme 2 (ACE2), which is found in several tissues, including the GI epithelial cells and liver cells (*4, 5*). SARS-CoV-2 was detected in biopsies of several tissues, including esophagus, stomach, duodenum and rectum, and endoscopy of hospitalized patients revealed esophageal bleeding with erosions and ulcers (*2, 6*).

Higher levels of ACE2 in the tissues may explain in part some of the comorbidities associated with severe COVID-19. Recently, we showed that *ACE2* was highly expressed in the lungs of people with pulmonary arterial hypertension and chronic obstructive diseases (*7*). Since the expression of *ACE2* changes under conditions of cell stress, elevated glucose levels and hypoxia (*8, 9*), other comorbidities related to the GI tract can be associated with different forms of COVID-19.

Here we suggest that gastroesophageal reflux disease (GERD) and Barrett’s esophagus (BE) may represent novel comorbidities associated with COVID-19. In the United States, it has been estimated that 5.6% of adults have BE, a disease where GERD damages the esophageal squamous mucosa (*10*). Here we demonstrated that *ACE2* is highly expressed in the esophagus of patients with BE and that the acid pH associated with this condition is a key inducer of *ACE2* expression. Human primary monocytes cultured in reduced pH display increased expression of ACE2 and increased viral load upon SARS-CoV-2 infection. We also show that patients using proton pump inhibitors, which are recommended for GERD treatment, have a higher risk of developing severe COVID-19, observed by an increased risk of ICU admittance and death.

## Methods

### Acidosis and Barrett’s esophagus meta-analysis

We manually curated the Gene Expression Omnibus (GEO) repository (https://www.ncbi.nlm.nih.gov/geo/) to find esophagus transcriptome datasets related to “Barrett’s esophagus” and cell line transcriptome datasets related to “acidosis” and “pH reduction”. Author-normalized expression values and metadata from these datasets were downloaded using the GEOquery package (*11*). We performed differential expression analyses using the limma package (*12*). The GEO study ID and the groups of samples compared are listed in Supplementary Table 1. The MetaVolcanoR package (*13*) was used to combine the P values using the Fisher’s method. To adjust for multiple comparisons, we calculated the false discovery rate (FDR) using the Benjamini-Hochberg procedure. For enrichment analyses, we utilized the EnrichR tool (*14*) and fgsea R package (*15*) with gene sets from the Gene Ontology Biological Process database. We then selected pathways with a P value adjusted for multiple comparisons lower than 0.10.

### Single cell transcriptomic analysis of Barrett’s esophagus

The single cell RNA-seq (scRNA-seq) data from esophagus, Barrett’s esophagus, gastric and duodenum cells from patients with BE were acquired from Owen et al. 2018 (*16*). Cells with less than 1,000 genes were excluded from analysis using Seurat v3 (*17*). Raw UMI counts were log transformed and variable genes called on each dataset independently based on the VST method. The *AddModuleScore* function was used to remove batch effects between samples and based on *C1orf43, CHMP2A, EMC7, GPI, PSMB2, PSMB4, RAB7A, REEP5, SNRPD3, VCP, VPS29* genes. We assigned scores for S and G2/M cell cycle phases based on previously defined gene sets using the *CellCycleScoring* function. Scaled z-scores for each gene were calculated using the *ScaleData* function and regressed against the number of UMIs per cell, mitochondrial RNA content, S phase score, G2/M phase score, and housekeeping score. Scaled data was used as an input into PCA based on variable genes. These PCA components were used to generate the UMAP reduction visualization. To identify the number of clusters, UMI log counts were used as input to SC3 (*18*). Technical variation was tested using BEARscc (*19*), which models technical noise from ERCC spike-in measurements. The clusters were then annotated based on genes previously characterized (*16*).

### Peripheral blood mononuclear cells (PBMC) isolation

Buffy coats provided by the Hematology and Hemotherapy Center of the University of Campinas (SP-Campinas, Brazil) were used for PBMC isolation as described (*9*). The study was approved by the Brazilian Committee for Ethics in Human Studies (CAAE: 31622420.0.0000.5404). Briefly, buffy coats were mixed and then diluted in Phosphate Buffer Saline (PBS) (1:1) and carefully to 50 mL tube containing Ficoll (Sigma-Aldrich) and centrifuged. PBMCs were cultured in RPMI 1640 for 2-3h to allow cell adhesion. Next, cells were washed twice with PBS and adherent cells, enriched in monocytes, were further incubated until infection in RPMI 1640 containing 10% fetal bovine serum (FBS) and 1% Penicillin-Streptomycin (Pen-Strep) at 37°C with 5% CO2. Monocytes were maintained in different pH levels (6, 6.5, and 7.4) during 24h and subsequently infected with SARS-CoV-2, as described below.

### Viruses and infection

HIAE-02 SARS-CoV-2/SP02/human/2020/BRA (GenBank MT126808.1) virus was isolated as described (*9*). Stocks of Sars-CoV-2 were prepared in the Vero cell line. The supernatant was harvested at 2–3 dpi. Viral titers were obtained by plaque assays on Vero cells. Monocytes were infected with SARS-CoV-2 at MOI 0.1 under continuous agitation at 15 rpm for 1 h. Next, monocytes were washed twice and incubated in RPMI with 10% FBS and 1% Pen-Strep for 24h at 37°C with 5% CO2 for 24 hours.

### Viral load and gene expression analyses

Total RNA extraction was performed using TRIzol Reagent (Sigma-Aldrich). RNA concentration was measured with NanoDrop 2000 spectrophotometer (Thermo Scientific). RNA was reverse-transcribed using GoScript™ Reverse Transcriptase cDNA synthesis kit following manufacturer’s instructions. SARS-CoV-2 viral load was determined with primers targeting the N1 region and a standard curve was generated as described (*20*). Viral load and gene expression were made using SYBR Green Supermix in BIO-RAD CFX394 Touch Real-Time PCR Detection System. Fold change was calculated as 2^^^-ΔΔCt. Primer sequences used: 18S (Forward: 5’-CCCAACTTCTTAGAGGGACAAG-3’; Reverse: 5’-CATCTAAGGGCATCACAGACC-3’); ACE2 (Forward: 5’-GGACCCAGGAAATGTTCAGA-3’; Reverse: 5’-GGCTGCAGAAAGTGACATGA-3’); SARS-CoV-2_IBS_N1 (Forward: 5’-CAATGCTGCAATCGTGCTAC-3’; Reverse: 5’-GTTGCGACTACGTGATGAGG-3’).

### Clinical data analysis

We retrieved clinical data from two independent cohorts of 551 and 806 RT-qPCR confirmed COVID-19 patients aged 18 years or older that went to reference hospitals for COVID-19 in Manaus, Amazonas, Brazil (North region cohort) and in São José do Rio Preto city, São Paulo, Brazil (Southeast region cohort), respectively. They were followed for at least 28 days (North region cohort) or 120 days (Southeast region cohort) after recruitment. Information about the previous history of proton pump inhibitors use (e.g. omeprazole and pantoprazole), a surrogate evidence of low gastric pH-related diseases, time of hospitalization, ICU admittance, and time to death, as well as demographics, previous use of other drugs, clinical, laboratory, and outcome variables were collected. The protocol was approved by the Brazilian Committee of Ethics in Human Research (CAAE: 30152620.1.0000.0005 and 30615920.2.0000.0005 for North region cohort, and 31588920.0.0000.5415 for Southeast region cohort). Data were collected and managed using REDCap (v. 10.2.1) electronic data capture tools hosted at *Fundação de Medicina Tropical Dr. Heitor Vieira Dourado*.

Adjusted hazard ratios and risk ratios with respective 95% confidence intervals (CI) were estimated for time to death and ICU admittance, respectively by Cox regression and log-binomial generalized linear model models. To adjust for confounders, ages higher than 60 years old and obesity, defined by both BMI and fat percentage, were used as covariables in the multivariable analyses. Wilcoxon Rank-Sum analysis was used to test differences in the days of hospitalization. A 2-tailed *P*<0.05 was considered significant. The statistical analyses were carried out using Stata v. 13.0 (StataCorp LP, College Station, TX).

## Results

To evaluate whether people with BE may have higher chances of being infected with SARS-CoV-2 when compared to people without the disease, we performed a meta-analysis of 8 transcriptomic studies of BE (Figure 1A, Table S1). A total of 304 and 256 genes displayed, respectively higher and lower expression BE compared to normal esophagus tissue in at least 7 of these studies (Figure 1B). *ACE2* was among the genes consistently up-regulated in the BE compared to normal esophagus (Figure 1C). While pathways related to keratinocyte differentiation and epidermis development were enriched with down-regulated genes, we found that bicarbonate transport and regulation of intracellular pH pathways were enriched with up-regulated genes (Figure 1D), suggesting that pH may influence ACE2 expression. In fact, when human coronary artery endothelial cells were treated with proton pump inhibitors – omeprazole or lansoprazole – the expression of ACE2 decreased in comparison to untreated cells (Figure 1E). Gene set enrichment analysis (GSEA) confirmed that Barrett’s esophagus tissues have higher expression of genes related to pH alterations (Figure 1F).

**Figure 1.**
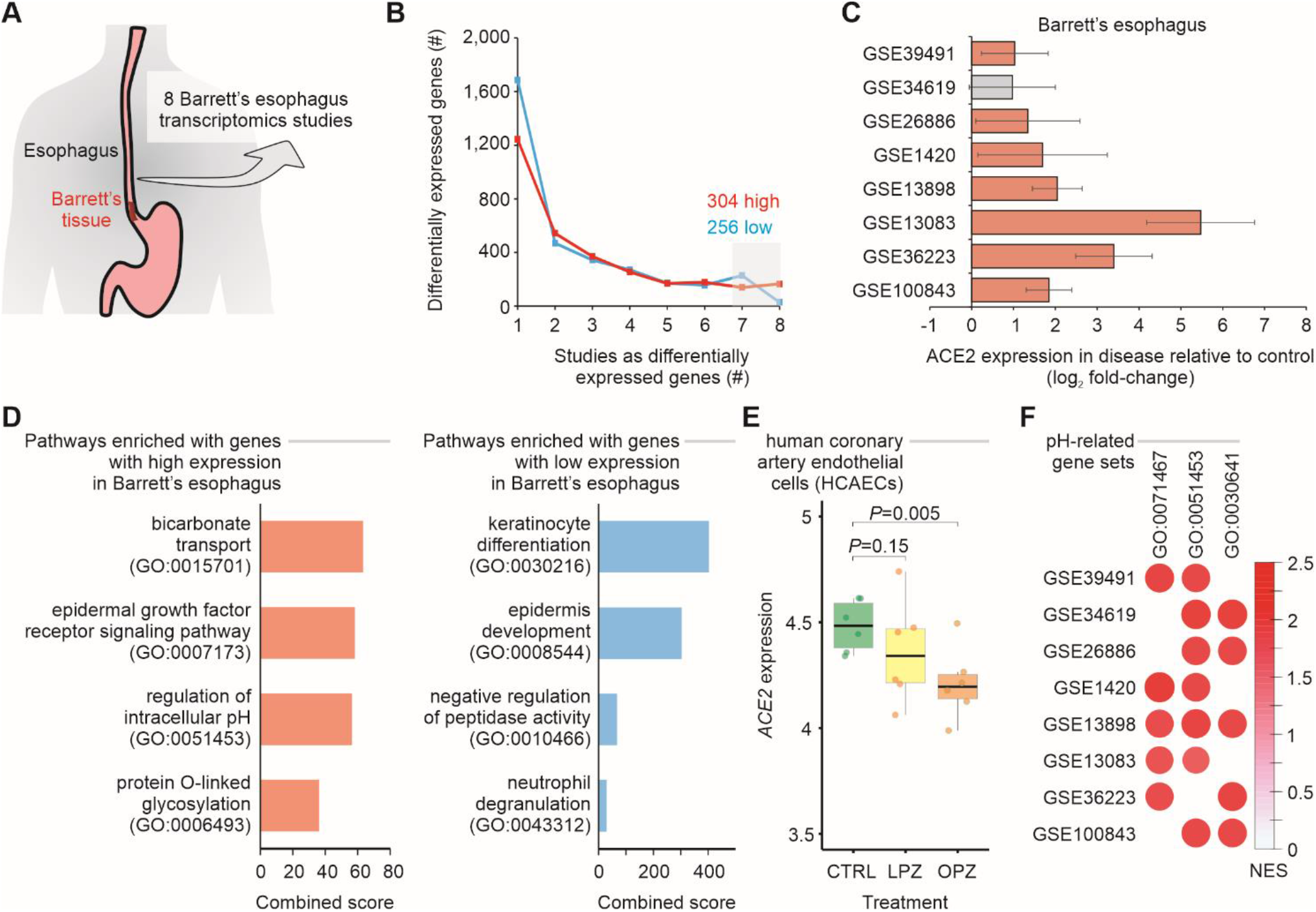
Meta-analysis of gastroesophageal junction transcriptomes of patients with Barrett’s esophagus. **A**. Meta-analysis of 8 studies of Barrett’s esophagus transcriptomes. **B**. Number of differentially expressed genes in Barrett’s esophagus compared with non-Barrett’s esophagus. The lines show the number of genes (y-axis) considered up-regulated (red lines) or down-regulated (blue lines) in Barrett’s esophagus (P value < 0.05; log2 fold-change > 1; combined FDR < 0.01) in one or more datasets (x-axis). The numbers of up-regulated and down-regulated genes in at least 7 studies are indicated. **C**. *ACE2* is upregulated in patients with Barrett’s esophagus. Each bar represents the log2 expression fold-change between patients and control individuals. The error bars indicate the 95% confidence interval. Bars in red represent a P value < 0.05 and in grey a non-significant P value. **D**. Pathway enrichment analysis using the up-regulated and down-regulated genes in at least 7 studies. The bars represent the combined score (x axis) calculated by Enrichr tool for selected Gene Ontology gene sets (y axis). **E**. ACE2 expression in cells treated with proton pump inhibitors. Each boxplot represents the log2 expression of untreated (CTRL) cells and cells treated with either omeprazole (OPZ) or lansoprazole (LPZ). **F**. Gene Set Enrichment Analysis (GSEA) of the 8 studies of Barrett’s esophagus transcriptomes using pH-related gene sets. The size and color of the circles are proportional to the normalized enrichment score (NES) of the gene sets (columns) on each study (rows). The Gene Ontology IDs are indicated at the top.

We also investigated *ACE2* expression in Barrett’s esophagus at single-cell level. Our analysis showed that single cells from Barrett’s esophagus patients are distinct than normal esophagus cells, as well as cells from duodenum and gastric tissues (Figure 2A). While a large fraction of duodenum cells expresses *ACE2* (*21*), only 11% of the single cells from Barrett’s samples have *ACE2* expression above 0 (Figure 2B). However, among the cells expressing *ACE2*, higher levels of the gene were found in gastric, Barrett’s, and duodenum cells when compared to esophagus cells (Figure 2C). Using GSEA, we found that genes associated with regulation of cellular pH were enriched among the up-regulated genes in gastric, Barrett’s and duodenum cells when compared to esophagus cells (Figure 2D).

**Figure 2.**
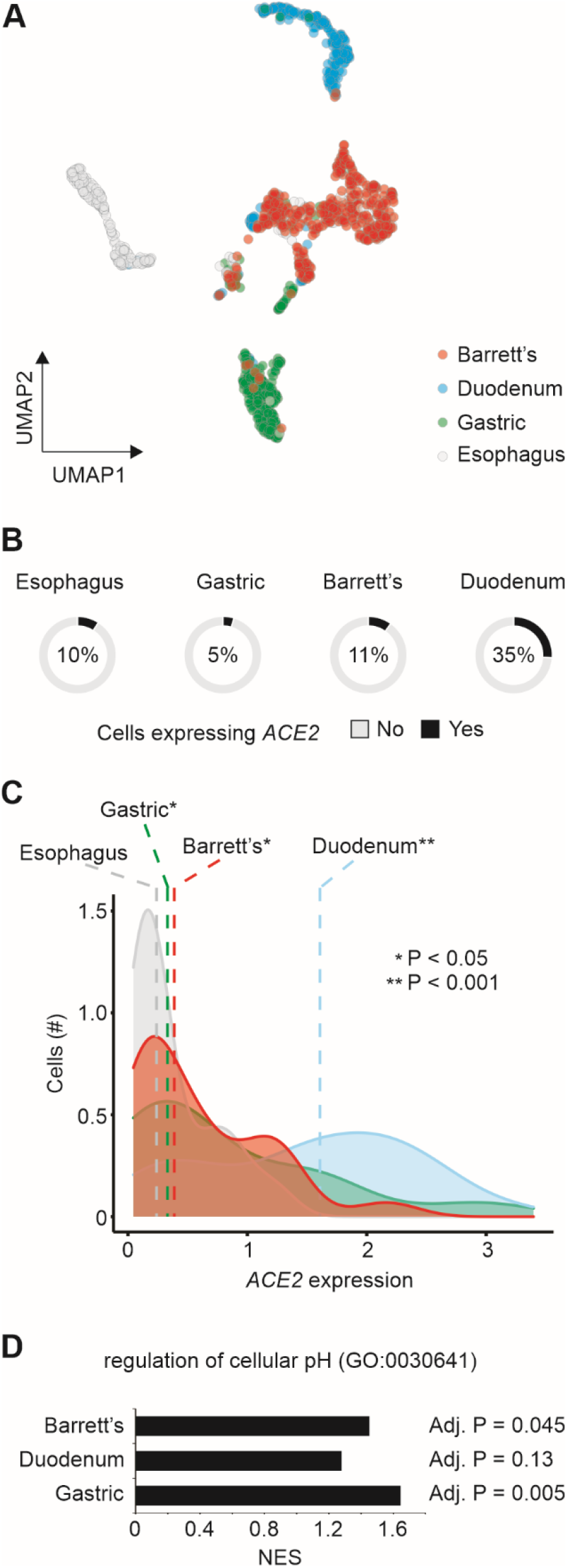
Single cell transcriptomics of Barrett’s esophagus. **A**. Dimension reduction of single cells using Uniform Manifold Approximation and Projection (UMAP). Cells from 4 patients with Barrett’s esophagus (n = 1,168) are shown. The colors represent the tissue types. **B**. ACE2 expression by tissue type. The pie charts show the number of single cells with (black) or without (grey) ACE2 expression (expression values > 0). The fractions of ACE2-expressing cells are indicated. **C**. Distribution of ACE2 expression by cells from different tissue types. The colors of histograms represent the tissue types. The dashed vertical line shows the median values of each tissue type. Student’s t-test P-value between tissue types versus esophagus is indicated. **D**. Gene Set Enrichment Analysis (GSEA) of the 3 tissue types compared to esophagus using the regulation of cellular pH gene set. The normalized enrichment score (NES) are shown in the x-axis for each one of the tissue types. The adjusted P-value of the enrichment is displayed right next to the corresponding bar.

To further evaluate whether pH may influence the expression of *ACE2*, we analyzed publicly available transcriptomic studies of cells under experimentally-induced acidosis. Cells cultured at lower pH displayed higher expression levels of *ACE2* when compared to those cultured under higher pH (Figure 3A and B). We validated this finding with human primary monocytes cultured at pH 7.4, 6.5 and 6.0 under normoxia. ACE2 expression was significantly increased at pH 6.5 and 6.0 compared to pH 7.4 (Figure 3C). The reduction of pH alone also significantly increased SARS-CoV-2 infection of human monocytes (Figure 3D), indicating that pH plays a role in ACE2-mediated SARS-CoV-2 infection.

**Figure 3.**
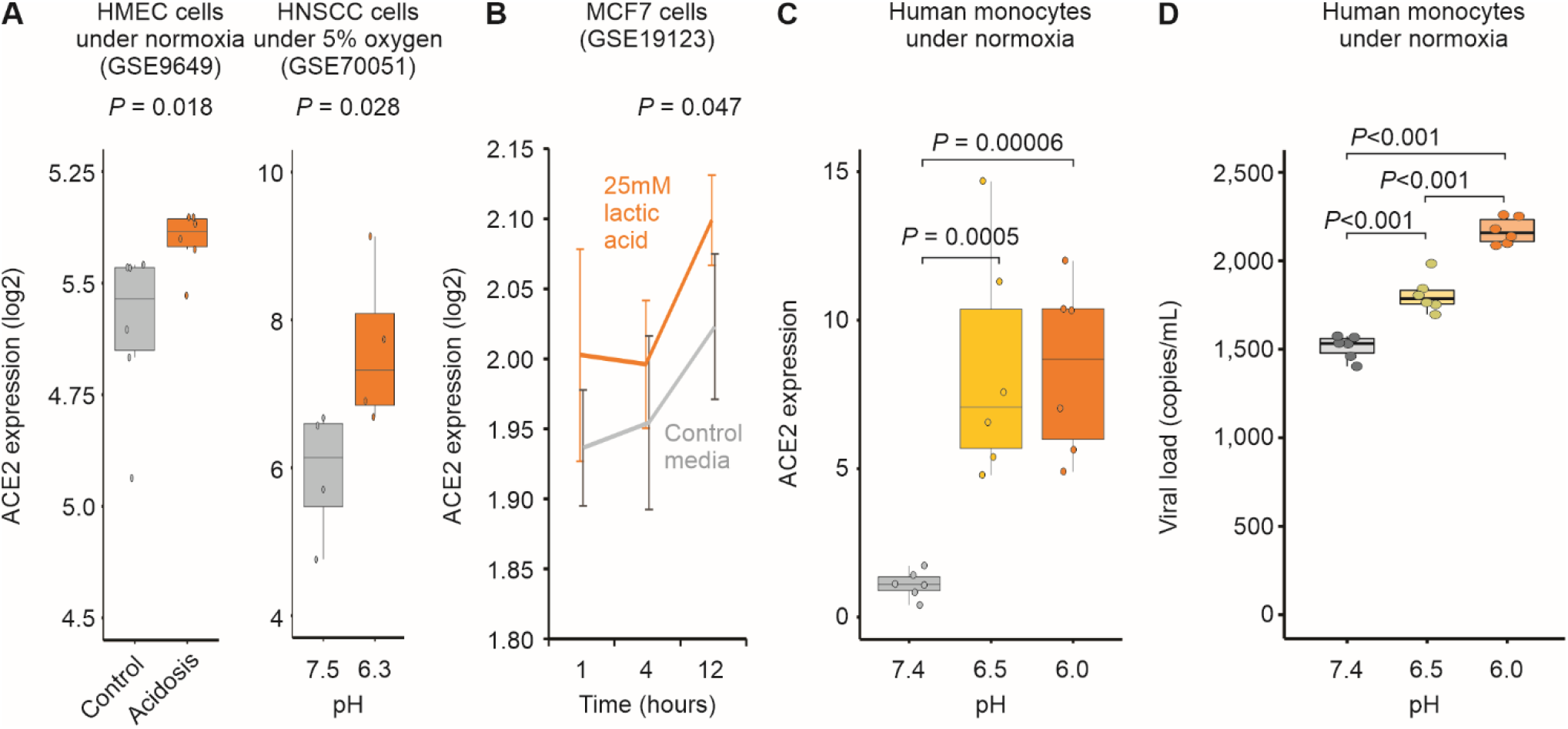
Acidosis increases ACE2 expression and SARS-CoV-2 infection. **A**. Human cells exposed to acidosis. Each boxplot represents the log2 expression of samples untreated (grey) or treated with lactic acidosis (brown) for two microarray studies (GSE9649 and GSE70051). Student’s t-test P-values are indicated. **B**. MCF7 cells exposed to pH reduction increases ACE2 expression. Grey and brown lines represent, respectively cells treated with control media or with 25mM lactic acid for 1, 4, and 12 hours (x-axis). Each point represents the mean log2 expression and the error bars the standard deviation of biological replicates. **C**. Acid pH increases ACE2 expression in monocytes. Human peripheral blood monocytes were incubated in medium at 3 different pH (6, 6.5, 7.4) for 24h. Each boxplot represents the fold change ACE2 expression. **D**. Acid pH increases SARS-CoV-2 viral load. Human peripheral blood monocytes were incubated in medium at 3 different pH (6, 6.5, 7.4) for 24h. The cells were infected with CoV-2 (MOI 0.1) for 1h under continuous agitation. The RNA viral load was measured by qPCR.

Proton pump inhibitors (PPI) decrease the amount of acid produced in the stomach and are often utilized to treat subjects with GERD symptoms or with certain stomach and esophagus problems (*22*). The use of PPIs prior to COVID-19 may serve as a proxy for identifying subjects with tissue irritation and inflammation caused by stomach acid. In two independent cohorts of 551 and 806 RT-qPCR confirmed COVID-19 patients from North and Southeast regions of Brazil, respectively, we investigated the effects of gastrointestinal discomfort and COVID-19 severity. Survival curve analysis showed that people using PPIs had a 2- to 3-fold higher risk of death compared to those not using the drug (Figure 4A). When controlling for potential confounders (i.e. age above 60 years old, diabetes, and hypertension), the adjusted hazard ratio was 2.183 (95CI: 1.635 - 2.914; P<0.0001) for the North region cohort and 2.332 (95CI: 1.661 - 3.274; P<0.0001) for the Southeast cohort (Figure 4B). These clinical findings indicate that the reduction of physiological pH (caused by stomach acid) may play a significant role in SARS-CoV-2 infection and COVID-19 severity.

**Figure 4.**
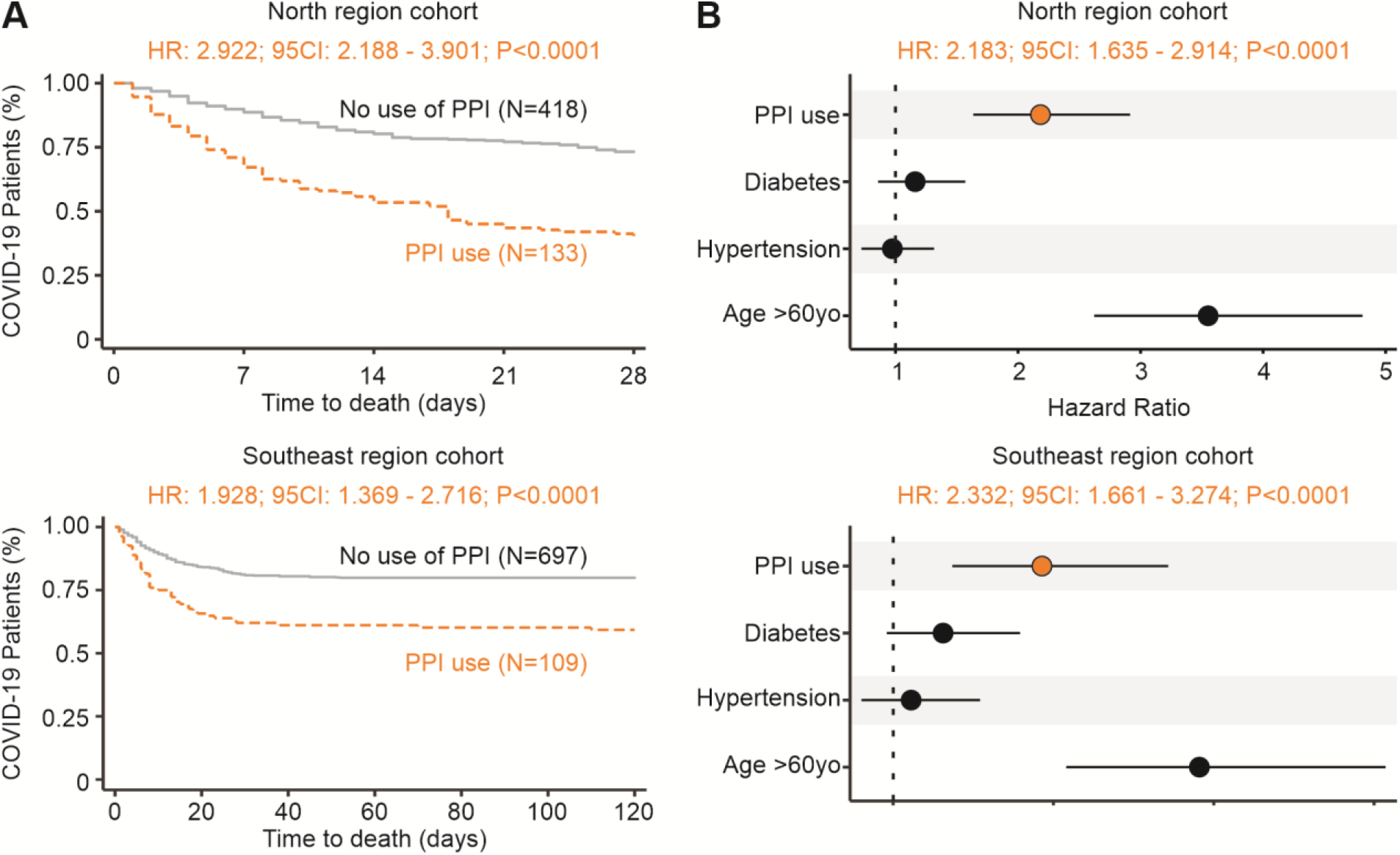
Increase risk of death in individuals with COVID-19 using proton pump inhibitors prior infection. **A**. Time to death. Kaplan-Meier survival curves showing a higher risk of death for the group of patients that used PPIs (brown) prior to admittance when compared to those not using them (grey). The North region cohort result is shown at the top and Southeast region cohort result is shown at the bottom. **B**. Risk of death. The forest plot presents the hazard ratios and respective 95CI for the main explanatory variable (brown), as well as the potential confounders (black) used in the multivariate model. The North region cohort result is shown at the top and Southeast region cohort result is shown at the bottom.

## Discussion

Our findings suggest that acid pH increases SARS-CoV-2 infection by up-regulating the ACE2 receptor, and this may have clinical implications for patients with GERD or Barrett’s esophagus. No clear mechanism exists linking alterations in pH and *ACE2* expression. Although evidence indicates that hypoxic conditions can increase the expression of *ACE2* (*8, 9*), the expression of neither SIRT1 nor HIF1A seem to be associated with Barrett’s esophagus (Table S2). We found that known regulators of ACE2 – HNF1B (*23*) and FOXA2 (*24*) – were up-regulated in 6 out of 8 Barrett’s esophagus transcriptomic studies (Table S2), suggesting that they may be involved with the pH-induced ACE2 expression in Barrett’s esophagus.

Pulmonary damage, one of the main features of severe COVID-19, may lead to acute hypoxia and further respiratory acidosis. It is possible that the acidosis in the blood of some patients with severe COVID-19 (*25*) worsen the disease by increasing the levels of ACE2 and facilitating the entry of SARS-CoV-2 into human cells. Hypoxia itself may contribute to the regulation of ACE2 (*9, 26*). In addition, elevated levels of the enzyme lactate dehydrogenase (which converts lactate from pyruvate) has been associated with worse outcomes in patients with COVID-19 (*27*). The excess of lactate may directly alter the extracellular and intracellular pH which in turn can impact ACE2 expression. The extent to which acute systemic acidosis contributes to COVID-19 severity is poorly known and deserves further research.

The drug famotidine suppresses gastric acid production by blocking the histamine 2 receptor in the stomach. Recently, Freedberg et al (*28*) have shown that early treatment of patients tested positive for SARS-CoV-2 significantly improved clinical outcomes among the hospitalized patients. Although the authors hypothesized that famotidine may have antiviral effects, it is possible that pH itself can play an important role in regulating ACE2 expression and limiting SARS-CoV-2 infection in patients.

We showed here that the previous use of PPIs is associated with unfavorable outcomes, such as the time of hospitalization, ICU admittance, and death. To the best of our knowledge, none of these associations were previously reported. Almario et al. (*29*) recently described that individuals using PPIs had higher chances for testing positive for COVID-19 when compared to those not using PPIs. Their hypothesis is that PPIs might increase the risk for COVID-19 by undermining the gastric barrier to SARS-CoV-2 and reducing the microbial diversity in the gut (*29*). Rather, we believe that PPIs are important markers of hidden comorbidities that involve the damage caused by the excess stomach acid in GI tissues.

By going from disease (Barrett’s esophagus) to molecule (ACE2) to cells (in vitro experiments) and back to clinical findings (COVID-19 patients), we showed that pH may have a great influence on SARS-CoV-2 infection and COVID-19 severity. Additional studies should be performed to not only confirm the clinical findings on a larger scale but also to assess the molecular mechanism related to pH-induced ACE2 expression.

## Data Availability

Publicly available transcriptomic data were obtained from the Gene Expression Omnibus (GEO) repository (https://www.ncbi.nlm.nih.gov/geo/)

## Funding

This work was supported by Brazilian National Council for Scientific and Technological Development (grant number 313662/2017-7); the São Paulo Research Foundation (grant numbers 2018/14933-2; 2018/21934–5; 2017/27131-9; 2013/08216-2; 2020/04836-0); and CAPES.

## Author declaration

Authors declare no competing interests.

## Author contributions

L.J., A.E.R.O., L.K.K.F., H.I.N. performed the transcriptome analyses. A.C.C., G.G.D., L.B.M., J.V.V, G.F.S., S.P.M., J.L.P., P.M.M. performed the experimental work. V.S.S., M.G.S.B., N.Z., F.A.G., M.L.N., F.A.V., G.C.M., W.M.M., M.V.G.L. performed the clinical analysis. H.I.N. coordinated the study. L.J. and H.I.N. wrote the manuscript with inputs from all of the co-authors.

## Supplementary Materials

Tables S1 and S2

## Notes

### Competing Interest Statement

The authors have declared no competing interest.

### Author Declarations

The protocol was approved by the Brazilian Committee of Ethics in Human Research (CAAE: 30152620.1.0000.0005 and 30615920.2.0000.0005).

